# Phenotype-by-phenome-wide association study of treatment resistant depression

**DOI:** 10.1101/2022.08.23.22279074

**Authors:** Brandon J Coombes, Jorge A Sanchez Ruiz, Brian Fennessy, Vanessa Pazdernik, Prakash Adekkanattu, Nicolas A Nunez, Lauren Lepow, Euijung Ryu, Ardesheer Talati, Greg D Jenkins, Richard Pendegraft, Priya Wickramaratne, J John Mann, Mark Olfson, Myrna M Weissman, Jyotishman Pathak, Alexander W Charney, Joanna M Biernacka

**Affiliations:** Department of Quantitative Health Sciences, Mayo Clinic, Rochester, MN; Department of Psychiatry & Psychology, Mayo Clinic, Rochester, MN; Department of Genetics and Genomic Sciences, Icahn School of Medicine at Mount Sinai, New York, NY; Department of Population Health Sciences, Weill Cornell Medicine, New York, NY; Clinical and Translational Science Center, Weill Cornell Medicine, New York, NY; Department of Psychiatry, Icahn School of Medicine at Mount Sinai, New York, NY; Department of Psychiatry, Columbia University & NY State Psychiatric Institute, New York, NY; Department of Psychiatry, Weill Cornell Medicine, New York, NY; Mount Sinai Clinical Intelligence Center, Icahn School of Medicine at Mount Sinai, New York, NY

## Abstract

**Objective:** Treatment-resistant depression (TRD), defined as inadequate response to at least one or at least two antidepressant (AD) trials, is common in major depressive disorder (MDD). In this study, electronic health records (EHR) were used to identify clinical associations with TRD.

**Methods:** Using two biobanks, phenomes of patients with at least one MDD-related diagnostic code and one AD prescription (N=17,049) were generated using aggregated diagnostic codes (phecodes) from EHRs. Phenotype-by-phenome-wide association analyses were performed for two binary definitions of TRD, based on either one or more, or two or more, AD switches after at least 30 days but within 14 weeks, and a quantitative measure defined as the number of unique ADs prescribed for at least 30 days.

**Results:** Of the 17,049 patients with MDD, 1624 (9.5%) had at least one switch, 422 (2.5%) had at least two switches, and the number of unique antidepressant prescriptions ranged from one to twelve. After accounting for multiple testing, 142, 18, and 7 phecodes were significantly associated with the quantitative definition and the two binary definitions (≥1 AD switch or ≥2 AD switches), respectively. All three outcomes were significantly associated with known TRD risk factors including anxiety disorders, insomnia, and suicidal ideation. The quantitative measure was uniquely associated with other conditions including irritable bowel syndrome and decreased white blood cell count.

**Conclusions:** In addition to identifying known clinical associations, the quantitative measure of treatment resistance uncovered new factors potentially associated with TRD. This measure may also facilitate discovery of genetic correlates of TRD in future analyses.

## Introduction

Major Depressive Disorder (MDD) is a heterogeneous disorder with a clinical presentation that varies significantly among individuals (1). Treatment selection for MDD is largely based on the expected tolerability of therapeutic interventions and patient-reported symptom severity. Two thirds of patients starting treatment for MDD in a large landmark naturalistic trial of standard treatment did not achieve remission on their first medication trial, and after four “adequate” interventions, only 67% remitted (2). Each successive therapeutic intervention involved a lower probability of achieving remission. Additionally, people with comorbid anxiety or anxiety disorders, obsessive-compulsive disorder, trauma-related disorders, melancholic features, or worse symptom severity were less likely to achieve remission (3). Non-remission has also been associated with having severe medical comorbidities or poor physical health, poor treatment adherence, current unemployment, and not having a college education (4, 5).

Treatment-resistant depression (TRD) does not have a singular consensus definition but is typically defined as an incomplete response to at least one or two antidepressant trials of adequate dose, duration, and adherence (6). Yet, such a dichotomous definition is an arbitrary distinction and does not capture the spectrum of resistance to treatment. A consequence is that studies using a dichotomous TRD definition may have suboptimal statistical power. To address this shortcoming, quantitative scoring of treatment resistance has been proposed based on variables such as the number and type of failed therapeutic trials or the use of complex interventions (e.g., electroconvulsive therapy), as well as depression severity measures (7).

Analysis of data from electronic health records (EHRs) can facilitate large scale clinical studies at a much lower cost than traditional medical research (8). In addition to the large and often diverse samples accessible through EHRs, EHR research also benefits from the wide range of data available including prescribed medications and diagnoses made by physicians allowing for comprehensive studies of comorbidities related to a disease or trait of interest. Aided by EHRs, phenotype-by-phenome-wide association studies (Phe^2^WAS) can identify associations between a phenotype that can be defined based on data in the EHR (such as measures of TRD) and a broad range of clinical diagnoses (i.e. the clinical “phenome” using groupings of International Classification of Diseases [ICD] codes) to further our understanding of comorbidities that are associated with the primary phenotype (9). In our study, we defined three measures of TRD from the EHR and performed a Phe^2^WAS for each TRD measure in two biobanks. We use the results to gain insight into possible phenotypic associations with TRD and to assess consistency of results and power across different measures of TRD.

## Methods

### Data sources

The data was derived from two biobanks: Mayo Clinic Biobank (MCB)(10) linked to the Mayo Clinic (Rochester, Minnesota and Jacksonville, Florida) and the Mayo Clinic Health System in La Crosse, Wisconsin and Bio*Me* linked to the Mount Sinai Health System (MSHS) in New York City. For the MCB, enrollment began in April 2009 and active enrollment ended in March 2016. Participants were aged 18 years and older and were selected largely through medical visits to primary care departments at the clinic. In total, nearly 60,000 participants were enrolled in the MCB. Bio*Me* began in September 2007 and recruited more than 60,000 individuals primarily in MSHS ambulatory care settings. At consent, both MCB and Bio*Me* participants provided biological samples, completed a questionnaire, and permitted researchers with approved access to search their full EHR from past and future clinical visits. The EHR data includes clinical notes, demographics, medications/prescriptions, laboratory values, billing codes from the ICD, 9^th^ and 10^th^ editions (ICD-9/10) (11), and Current Procedural Terminology (CPT) codes. We mapped ICD9/10 codes to phecodes (i.e., higher order groups of diagnoses) as described and validated by the Phecode map 1.2b1 (9). The current study was reviewed and approved by each site’s Institutional Review Board and exempted from informed consent requirements as non-human subjects research (Mayo Clinic IRB approval 19-006227 and Mount Sinai IRB approval 07-0529). For the current study, EHRs for MCB and Bio*Me* participants were extracted in September and December, respectively, in 2021.

### Defining cases with MDD

Using structured EHR data, MDD was defined as having at least one MDD-related ICD9/10 code, using an initial list of codes mapped to phecodes for MDD (phecodes 296.2 and 296.22), available from https://phewascatalog.org/phecodes (12) with the addition of dysthymic disorder (ICD9:300.4; ICD10:F34.1), depressive type psychosis (ICD9:298.0), and atypical depressive disorder (ICD9:296.82) (Supplementary Table 1). Participants with phecodes for bipolar disorder (phecode 296.1) or psychotic disorders (phecodes 295.1, 295.2, 295.3, 295) were excluded from the MDD samples (Supplementary Table 2).

### Prescription data

For the Mayo Clinic sample, outpatient drug information was extracted from prescription data using Mayo Clinic’s open-source cTAKES natural language processing (NLP) platform and mapping to RxNorm codes (https://www.nlm.nih.gov/research/umls/rxnorm) (13). The process of retrieving prescription data for Bio*Me* has been previously described (14). Briefly, drug prescription data were obtained from the Mount Sinai Data Warehouse (MSDW) and standardized to RxNorm concept unique identifiers (RXCUIs) through an open source CLAMP clinical NLP pipeline (15) and to further standardize, RXCUIs were mapped to base ingredient information using the RxNorm application program interface.

In both samples, we abstracted prescription data for the antidepressants listed in Supplementary Table 3. Notably, we excluded clomipramine, fluvoxamine, milnacipran, selegiline, and trazodone because, while in the antidepressant drug class, these drugs are commonly used to treat other neuropsychiatric symptoms and disorders. Patients without prescription data were removed from the analysis.

### Algorithm for Defining TRD

As described above, participants with MDD were identified as those having at least one code for a depressive disorder and no diagnostic codes for bipolar disorder or psychotic disorders. Within this set of participants, we restricted the sample to those with at least one antidepressant prescription in their record.

For each antidepressant (listed in Supplementary Table 3), we retrieved the start and end dates (i.e., after refills have run out) and calculated the total time potentially on the medication. If another medication from the list was prescribed before the prescription end date, the duration on the given medication was truncated to end at the start of the new prescription to indicate a change in antidepressant treatment. To determine “adequate trials,” an antidepressant was excluded from the patient’s data if they switched to a new medication within 30 days of starting the antidepressant or were not prescribed the medication for at least 30 days on the antidepressant. This approach was taken to avoid capture of medications stopped due to intolerable side-effects rather than lack of efficacy. Finally, similar to a recent study by Fabbri and colleagues (16), a medication “switch” was defined if the time interval between the prescription of two consecutive drugs was no longer than 14 weeks. The resulting medication trial and switch data were then used to define one quantitative measure and two dichotomous measures of TRD.

#### Total number of unique antidepressants

The quantitative measure of TRD was defined as the number of unique antidepressants (ADs) that a participant was prescribed for at least 30 days.

#### Antidepressant switches

We also defined TRD using the standard dichotomous definitions of either one or more AD switches, or two or more AD switches.

#### Phe^2^WAS

To assess clinical conditions (i.e., diagnoses) associated with TRD in each biobank, we performed a Phe^2^WAS using each TRD outcome as the predictor of each phecode in a logistic regression model using the PheWAS R package (9). At least two occurrences of a diagnostic code in the EHR on different days were required to define a phecode case. For the Phe^2^WAS, we restricted tests of association to phecodes with a minimum of 50 cases and 50 controls. To account for potential confounders of EHR analyses such as healthcare utilization and demographics, we adjusted for the patient’s length of EHR (defined as the time between the first and last ICD code in the record), total number of ICD codes (across all conditions, not limited to those in Supplementary Tables 1 and 2), median age of the record (defined by taking the median of time since each ICD code in a patient’s record) as well as the patient’s age at time of the EHR data pull and self-reported race, ethnicity, and gender. Results from the two biobanks were meta-analyzed using a fixed-effects meta-analysis with the meta R package (17). To account for multiple testing, we used a Bonferroni corrected threshold that adjusted for the total number of tests across the three Phe^2^WAS (p < 0.05/[862 phecodes x 3 outcomes] = 2e-6). All analyses were performed in R 4.0.3.

## Results

Table 1 describes the MDD patient samples with at least one antidepressant medication treatment in the two biobanks (MCB N = 12944; Bio*Me* N = 4055). At both sites, the patient population majority was female (72% and 69%, respectively) with a median age of 67 years. Participants from the MCB were predominantly White reflecting the population around Rochester, Minnesota, whereas Bio*Me* had enrolled roughly equal proportions of White and Black participants and about half of the individuals were Hispanic, reflecting the neighborhoods surrounding Mount Sinai in New York City. Thus, among the participants with diagnosed MDD, the 90% of MCB sample was White while Bio*Me* was more diverse (25% White, 24% Black, and 49% Hispanic). Supplementary Tables 4 and 5 provide additional descriptive information about each of the biobanks.

**Table 1.** Description of the two biobank samples

| **Characteristic** |  | **MCB**<br>**(N=12994)** | **BioMe**<br>**(N=4055)** |
| --- | --- | --- | --- |
| Age, y | Median (Q1, Q3) | 67 (56, 78) | 67 (58, 77) |
| Female | N (%) | 9358 (72.0%) | 2793 (68.9%) |
| **Race** | N (%) |  |  |
| White |  | 11751 (90.4%) | 1008 (24.9%) |
| Black/African American |  | 97 (0.7%) | 963 (23.7%) |
| Asian |  | 75 (0.6%) | 36 (0.9%) |
| Native American/<br>Alaskan Native |  | 26 (0.2%) | 1 (0.02%) |
| Other |  | 65 (0.5%) | 1762 (43.5%) |
| Mixed/Unknown |  | 980 (7.5%) | 285 (7.0%) |
| Hispanic | N (%) | 198 (1.5%) | 1997 (49.2%) |
| **EHR measures** |  |  |  |
| Length of record, y | Median (Q1, Q3) | 21 (12, 32) | 11 (8, 13) |
| Median age of record, y | Median (Q1, Q3) | 8 (6, 11) | 9 (7, 10) |
| Log No. Diagnoses | Median (Q1, Q3) | 6.05 (5.41, 6.64) | 5.89 (5.12, 6.51) |
| **TRD measures** |  |  |  |
| No. unique ADs | Mean (Range) | 2.20 (1 to 12) | 2.07 (1 to 12) |
| 1+ AD switch | N (%) | 1252 (9.6%) | 372 (9.2%) |
| 2+ AD switch | N (%) | 372 (2.9%) | 50 (1.2%) |

AD: antidepressant; EHR: electronic health record; Age: age at date of data pull; Median age of record is calculated for each patient as the median time between each diagnosis date and date of data pull; Log No. diagnoses are the natural log of total number of ICD codes per patients

The distributions of the three TRD variables were similar across the two biobanks with patients receiving on average two different antidepressant prescriptions, and about 9% (N=1624) of the MDD sample having one or more antidepressant switches and 2% having two or more switches (Table 1). The meta-analysis Phe^2^WAS identified significant associations for all three measures of TRD (Figure 1). The full results including biobank-specific results are shown in Supplemental Table 6. Overall, after accounting for multiple comparisons across the three TRD measures, the quantitative measure of TRD had the most statistically significant associations with 142 phecodes associated with the number of ADs prescribed, followed by 18 phecode associations for the ≥1 AD switch and 7 for the ≥2 AD switches TRD measures.

**Figure 1.**
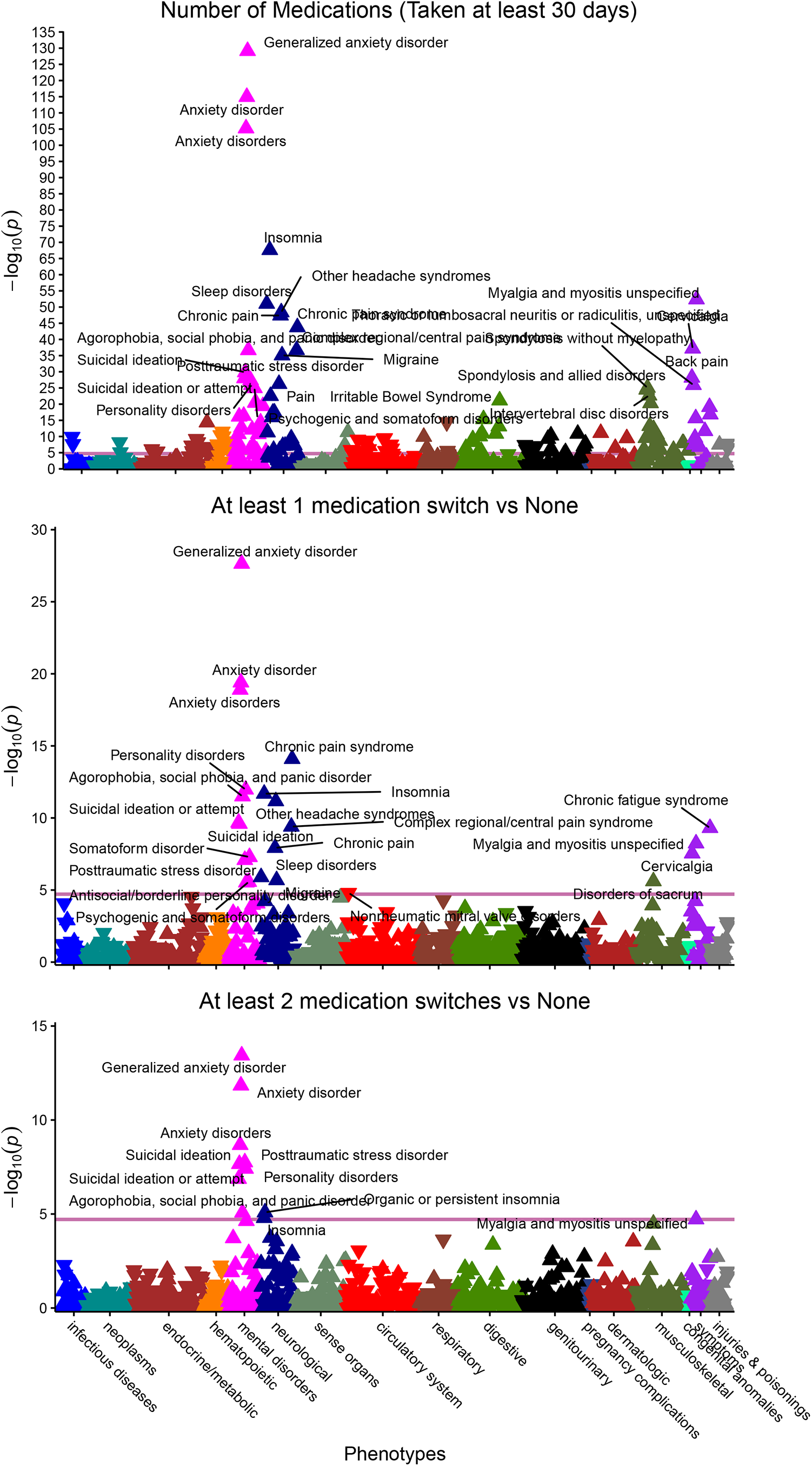
Association plot of Phe^2^WAS testing for association across the EHR phenome with three definitions of treatment resistant depression (TRD): (A) Number of unique prescriptions, (B) At least one switch of prescription within 14 weeks of previous prescription, (C) At least two switches of prescription within 14 weeks of previous prescription

Notably, we found consistent significant associations across the three measures of TRD including with suicidal ideation (phecode 297.1; number of ADs, OR = 1.40 per increase in AD prescribed, p = 1E-30; ≥1 AD switches, OR = 2.25, p = 3E-10; ≥2 AD switches, OR = 3.64, p = 2E-08), anxiety disorders (phecode 300.1; number of ADs, OR = 1.41, p = 1E-115; ≥1 AD switches, OR = 1.75, p = 4E-20; ≥2 AD switches, OR = 4.20, p = 1E-12), posttraumatic stress disorder (PTSD, phecode 300.9; number of ADs, OR = 1.29, p = 4E-28; ≥1 AD switches, OR = 1.72, p = 8E-08; ≥2 AD switches, OR = 2.92, p = 2E-08), personality disorder (phecode 301; number of ADs, OR = 1.31, p = 1E-26; ≥1 AD switches, OR = 2.15, p = 1E-12; ≥2 AD switches, OR = 3.02, p = 4E-08), insomnia (phecode 327.4; number of ADs, OR = 1.27, p = 2E-68; ≥1 AD switches, OR = 1.51, p = 2E-12; ≥2 AD switches, OR = 1.90, p = 2E-05), as well as myalgia and myositis (phecode 770; number of ADs, OR = 1.25, p = 4E-53; ≥1 AD switches, OR = 1.45, p = 6E-09; ≥2 AD switches, OR = 1.96, p = 2E-05).

Many phecodes (87% of the 142 significant associations) were only significantly associated with the quantitative measure of TRD, i.e. the number of ADs ever taken. Notably, irritable bowel syndrome (phecode 564.1; number of ADs, OR = 1.19, p = 7E-22; ≥1 AD switches, OR = 1.20, p = 0.03; ≥2 AD switches, OR = 1.57, p = 0.02), and decreased white blood cell count (phecode 288.1; number of ADs, OR = 0.84, p = 3E-9; ≥1 AD switches, OR = 0.63, p = 0.001; ≥2 AD switches, OR = 0.51, p = 0.09) were associated with number of ADs ever taken, but did not reach Bonferroni-corrected statistical significance thresholds for the two dichotomous measures of TRD.

## Discussion

Using data from two large biorepositories, we conducted a phenotype-by-phenome association study to test for associations of diagnostic phecodes with three different EHR-based measures of treatment-resistant depression. All three measures of TRD showed consistent associations with anxiety disorders, insomnia, myalgia and myositis, suicidal ideation or attempt, PTSD, and personality disorders, all of which have been previously shown to be associated with TRD. Notably, the strength of evidence for these associations was much higher when using the total number of unique prescribed antidepressants as the measure of TRD, suggesting that this quantitative measure provided greater statistical power. This quantitative TRD definition resulted in the largest number of significant associations with distinct clinical phenotypes including several that had not been previously shown to be associated with TRD, such as irritable bowel syndrome and decreased white blood cell count.

Our agnostic search for association with TRD across EHR diagnoses identified clinical phenotypes that have been previously reported to be associated with TRD such as anxiety, suicidal ideation, insomnia, PTSD, personality disorders, and pain. Mounting evidence indicates that having psychiatric or medical comorbidities is linked to worse treatment outcomes for people with MDD (18). In our study, a diagnosis of any anxiety disorder was the phenotype group that was most strongly associated with all measures of TRD, which is consistent with prior research (19–21). More specifically, generalized anxiety disorder, which is also commonly treated with antidepressants, had the strongest association among all anxiety disorders including agoraphobia, social phobia, and panic disorder, as previously described (22, 23). As expected, suicidal ideation, which has been consistently associated with TRD in the literature and a symptom of severity (19, 23–26), was strongly associated with TRD. Insomnia and suicide, both symptoms of and risk factors for MDD (27, 28), were associated with all measures of TRD. Interestingly, it was second only to anxiety disorders in the strength of its association with the total number of unique antidepressants. PTSD, which was significantly associated with all TRD outcomes, has also been associated with TRD in other studies (4, 25). This relationship could be mediated by past history of trauma, which results in increased severity of depression (29), which is in turn a risk factor for TRD (19, 21, 24), although evidence is conflicting (30). Finally, severe pain or lack of improvement of pain are known independent risk factors for TRD, regardless of the root cause (20).

Searching across the EHR also allowed us to make new discoveries of clinical phenotypes associated with TRD. Many clinical phenotypes were significantly associated with the three measures of TRD—especially with the total unique number of antidepressants tried. For example, the total unique number of antidepressants was associated with functional digestive disorders, including irritable bowel syndrome (IBS) and functional dyspepsia. People with functional gastrointestinal disorders have higher somatization, interpersonal sensitivity, and life event stresses (31) and those with IBS have increased rates of mood and anxiety disorders (32, 33). It has also been shown that mood and anxiety disorders may share some of the same genetic pathways with IBS (34) and IBS is sometimes treated with antidepressants as well (35). It is worth noting that one of the most common side effects of antidepressants is gastrointestinal issues and this finding could indicate trans-diagnostic somatic symptoms.

The quantitative measure of treatment resistance was also inversely associated with decreased white blood cell (WBC) count. However, because of the low prevalence of this phecode (∼4%), its association with the binary TRD outcomes was not statistically significant though the direction of effect was the same. A TRD association with WBC count potentially supports the neuroinflammation and stress response models of depression, which hypothesize an activated immune system contributes to risk of depression and that stress from depressive symptoms leads to a proinflammatory state (36). It is important to note though that some medications may lower WBC count and thus this finding may be related to polypharmacy effects. Interestingly, a recent EHR study also found that increased genetic risk of depression, which has been hypothesized to be associated with higher risk of TRD, was also associated with higher white blood cell count (37).

Among the three measures of TRD that we considered, the total number of unique antidepressants is noteworthy for being quantitative—rather than the historically used binary TRD outcome (38). The quantitative trait attempts to place treatment resistance on a continuum, more closely resembling what is seen in clinical practice. In addition, the total number of unique antidepressants prescribed is easily assessed with EHRs. The results we observed when using the quantitative definition of TRD were not only consistent with the binary definitions, but also revealed greater power to detect significant associations across the phenome. Prior studies of TRD that used prescription data from EHRs have focused on binary measures of TRD (16, 39) and did not agnostically test for clinical associations with TRD, but found TRD was associated with psychiatric and non-psychiatric comorbidities (particularly anxiety disorders). While our agnostic Phe^2^WAS was well-powered, the increase in statistical power provided by the quantitative outcome will be especially beneficial for genomic studies of TRD, which have thus far been under-powered (16).

Our study had several limitations. First, clinical phenotypes were derived solely from structured EHR data. Diagnostic codes alone may not capture the nuances of the clinical presentations of every illness, nor do they replace standardized diagnostic assessments. However, phecode groupings help generate a phenome that more closely resembles natural-language medical records using structured EHR data and may facilitate replication of known associations more closely than other diagnostic codes or groupings (12). Second, we recognize that counting the number of unique antidepressants for a patient may not truly reflect TRD and could be confounded by other illnesses that are treated with antidepressants including anxiety, pain syndromes, or sleep disorders which were some of the diseases most strongly associated with the total number of unique antidepressants in our analyses. Nonetheless, the high consistency between our findings and associations with other measures of TRD provides evidence to support our results. We took steps to mitigate the influence of higher healthcare utilization on the total number of antidepressants by adjusting for the number of unique diagnostic codes present in an EHR, the length of each participants’ medical record, and excluding antidepressants that are not routinely prescribed for MDD. In addition, we aimed to limit the impact that antidepressant tolerability may have had on the number of antidepressants exposures by including only prescriptions that lasted at least 30 days, as the efficacy of antidepressants might not be fully assessable within such a time frame, and early interruption would most likely be related to treatment intolerance or other factors. Third, EHR prescription data does not capture whether prescriptions are actually filled, making it challenging to assess adequacy of dose and impossible to assess adherence, both of which are important for accurately assessing efficacy of an antidepressant. Fourth, sampling of patients by biobanks in particular healthcare systems may limit external generalizability (40). Encouragingly, even though the two biobanks included in the analysis differ geographically and by race/ethnicity, the distributions of the three TRD measures were very similar between the two biobanks. Fifth, we defined MDD in our population of interest only based on ICD codes, which may have limited reliability. However, the study by Fabbri et al. (2021) found significant overlap between EHR-defined MDD using at least two depressive disorder diagnostic codes and different MDD definitions. After cross-validating EHR-defined MDD with primary care data, they highlighted that using a less restrictive definition (i.e., at least one diagnostic code) resulted in sufficient diagnostic overlap and similar associations with MDD polygenic risk score, making this a powerful yet still reliable source of case status for MDD. Finally, it is important to recognize that there are many correlations among phecodes and thus the associations from the Phe^2^WASs are not independent and must be interpreted accordingly.

Despite the limitations, a key strength to the large-scale EHR-based agnostic phenome-wide approach is the possible discovery of a broad range of clinical associations not limited to known or hypothesized ones. Discoveries made through this approach may then be explored with targeted studies. Furthermore, unlike diagnoses derived from EHRs, epidemiological studies of risk factors of psychiatric disorders often do not have medical diagnoses made by physicians nor the details of clinical information including all prescribed medications in a defined time period. Thus, EHR-based observational studies are complementary to traditional clinical research study designs and provide a unique opportunity for discovery of association patterns in a clinical context.

## Conclusion

Our study replicated many known associated clinical risk factors for TRD such as anxiety, suicidality, and insomnia, and identified new factors such as irritable bowel syndrome and abnormal white blood cell count. Results from the analysis of the quantitative measure of treatment resistance were consistent with, and yet more powerful, than results from analyses using the binary TRD outcomes. The superior statistical power of the quantitative measure may aid the detection of genetic correlates of TRD in future analyses, and might prove useful in the rapid clinical assessment of treatment resistance.

## Data Availability

All data produced in the present study are only available after institutional review of request of access to the electronic health record.

## Acknowledgements

This study was supported by the National Institute of Mental Health (NIMH) grants: R01MH121924, R01MH121923, R01MH121922, and R01MH121921.

## Notes

**Disclosures:** In the last three years, Dr. Weissman has received research funds from NIMH, Templeton Foundation, Brain and Behavior and the Sackler Foundation and has received royalties for publications of books on interpersonal psychotherapy from Perseus Press, Oxford University Press, on other topics from the American Psychiatric Association Press and royalties on the social adjustment scale from MultiHealth Systems. None of these represent a conflict of interest and all other authors have no other disclosures

### Competing Interest Statement

In the last three years, Dr. Weissman has received research funds from NIMH, Templeton Foundation, Brain and Behavior and the Sackler Foundation and has received royalties for publications of books on interpersonal psychotherapy from Perseus Press, Oxford University Press, on other topics from the American Psychiatric Association Press and royalties on the social adjustment scale from MultiHealth Systems. None of these represent a conflict of interest and all other authors have no other disclosures

### Author Declarations

The current study was reviewed and approved by each site's Institutional Review Board and exempted from informed consent requirements as non-human subjects research: Mayo Clinic IRB approval 19-006227 and Mount Sinai IRB approval 07-0529.

